# Evolving Ocular Safety Signals of EGFR Inhibitors: A FAERS Disproportionality Analysis of Amivantamab, Mobocertinib, and Classic Agents

**DOI:** 10.64898/2026.03.16.26348536

**Authors:** Zaid Khan, Arnie Nadel, Thomas J. Joly

## Abstract

**Background:** Epidermal Growth Factor Receptor (EGFR) inhibitors, while effective in oncology, are associated with under-characterized ocular adverse events (AEs). Prior studies have been limited in scope, lacking a comprehensive, class-wide analysis of the full spectrum of ocular toxicity, particularly for newer agents.

**Methods:** We conducted a disproportionality analysis of the FDA Adverse Event Reporting System (FAERS) (2001-2025). Twelve EGFR-targeted agents were evaluated against a pre-specified set of ocular MedDRA Preferred Terms. To ensure robust signal detection, a significant association was defined by ≥3 co-reported cases, a Proportional Reporting Ratio (PRR) ≥2.0, and a false-discovery-rate adjusted p-value <0.05.

**Results:** Among 6,976,462 drug-event combinations, 20 met all signal criteria for Eyelash Abnormalities, Ocular Surface Disease, or Vision-Threatening and Intraocular Events. Trichomegaly demonstrated extreme disproportionality (e.g., panitumumab PRR= 465.3, 95% Confidence Interval [CI], 247.7-874.3). A consistent pattern of ocular surface toxicity (conjunctivitis, keratitis, blepharitis) was observed across multiple tyrosine kinase inhibitors and monoclonal antibodies, indicating a class-wide effect. Signals for serious events included corneal perforation (erlotinib, n= 7, PRR=13.9, 95% CI= 6.6-29.4) and optic neuropathy (erlotinib, n= 6, PRR= 2.9, 95% CI= 1.3-6.4).

**Conclusion:** This analysis confirms a strong, class-wide signal for ocular toxicity across the spectrum of EGFR inhibitors, from characteristic eyelid changes to sight-threatening complications. These findings underscore the necessity for proactive ophthalmologic monitoring, including baseline assessment, in patients receiving these therapies to preserve vision and maintain quality of life during cancer treatment.

## Introduction

Epidermal growth factor receptor (EGFR) inhibitors have revolutionized the treatment landscape for various malignancies, especially non-small cell lung cancer, by offering targeted therapy with improved survival outcomes [1, 2, 3]. These agents include first-generation tyrosine kinase inhibitors (TKI) such as gefitinib and erlotinib, second-generation TKIs like afatinib, and third-generation compounds such as osimertinib, as well as monoclonal antibodies targeting the EGFR such as cetuximab. They function by directly blocking the receptor’s activation or by preventing EGFR dimerization and downstream TK mediated signaling pathways essential for cellular proliferation, differentiation, and migration [3, 4]. While generally better tolerated than conventional chemotherapy, EGFR inhibitors are associated with a characteristic profile of adverse effects, most notably dermatologic manifestations such as an acneiform rash [3, 5, 6].

Ocular toxicities represent a significant, though perhaps underrecognized, class of adverse events associated with EGFR inhibitor therapy. Early case reports and reviews from the mid-2000s first delineated the spectrum of these ocular effects, which can occur in approximately one-third of patients [6]. The manifestations are diverse, broadly affecting the ocular surface and adnexa. Common findings include symptomatic external ocular changes such as meibomitis, squamous blepharitis, tear film dysfunction leading to dry eye disease, periocular skin rash, and the phenomenon of trichomegaly (hyperpigmented, tortuous eyelash growth) [6, 7]. Other ocular complications have also been reported including conjunctivitis, superficial keratitis, and persistent corneal epithelial defects that can predispose to secondary infectious keratitis [8, 9, 10, 11, 12].

The biological plausibility of these findings is supported by the critical role of EGFR in ocular homeostasis. EGFR is constitutively expressed in the basal epithelial cells of the cornea, conjunctiva, and eyelid structures, where it drives the proliferation and stratification essential for wound healing [4, 13, 14, 15, 16]. Pharmacologic inhibition disrupts these downstream signaling cascades, impairing epithelial repair and barrier function. Clinically, this manifests as keratitis, persistent epithelial defects, and potential corneal perforation [8, 9, 10, 14]. Furthermore, EGFR signaling regulates the growth phase of the hair follicle cycle [17]. Blockade of this pathway perturbs follicular differentiation, resulting in the trichomegaly and eyelash abnormalities frequently observed with these therapies [6, 7, 18, 19].

In recent years, the understanding of EGFR inhibitor-associated ocular toxicity has been refined through large-scale pharmacoepidemiologic studies. Analyses of major pharmacovigilance databases, including the US FDA Adverse Event Reporting System (FAERS), have provided robust signal detection, confirming significant associations between EGFR inhibitors and various ocular disorders [20, 21]. Notably, recent cohort studies have specifically quantified an increased risk of keratitis among patients treated with EGFR inhibitors [8, 9]. However, many prior pharmacovigilance studies have focused on a subset of agents (e.g., only TKIs) or a limited range of ocular events, and to our knowledge, a comprehensive, class-wide comparison inclusive of the latest biologic agents is lacking [20, 22, 23].

This gap in the pharmacovigilance literature is critical, as the rapid evolution of cancer therapeutics – including the introduction of newer-generation EGFR inhibitors and their use in combination therapies – necessitates an updated safety profile. While prospective cohort studies provide essential incidence data, disproportionality analysis of spontaneous reporting systems like FAERS remains a powerful tool for the initial detection and hypothesis-generating characterization of potential adverse drug reactions, especially for rare or wide-ranging events across a large drug class. Therefore, to address this need, we conducted a comprehensive disproportionality analysis of the FAERS database.

The primary objective was to detect and characterize potential safety signals (i.e., indications of concern warranting further study) for a broad, pre-specified spectrum of ocular adverse events across the entire contemporary class of EGFR-targeted agents, providing a quantitative comparison of agent-specific risks. To enhance the rigor of our signal detection in the context of multiple comparisons, we employed a false discovery rate (FDR) correction, a methodological layer to reduce false-positives (Type 1 Errors). By focusing on disproportionality signals, this analysis seeks to inform clinical vigilance and prioritize hypotheses for future prospective studies aimed at determining true incidence and causality.

## Methods

### 2.1 Data Source and Extraction

We conducted a post-marketing pharmacovigilance study using the US Food and Drug Administration Adverse Event Reporting System (FAERS), a spontaneous reporting database containing de-identified safety reports from healthcare professionals, manufacturers, and consumers. Data was accessed via the public openFDA API [24]. Our analytical approach aligns with the FDA’s framework for data mining spontaneous reporting systems for safety signal detection [25]. All data extraction and analysis were performed using R (version 2025.05.1+513) in RStudio. The dataset included all reports from January 1, 2001, through August 31, 2025, queried on November 22, 2025. Each FAERS case was treated as unique at the level of the [`safetyreportid`] identifier as provided by openFDA. Of note, we did not attempt additional manual de-duplication beyond this identifier; the implications of potential residual duplication are addressed in the limitations section of the Discussion.

### 2.2 Exposure Definition: EGFR-Targeted Agents

We predefined a list of systemic anticancer agents with primary or secondary EGFR-inhibitory activity. The exposure variable was the generic drug name as recorded in the FAERS [**patient.drug.medicinalproduct]** field. All reports in which an EGFR inhibitor appeared in the patient.drug dataset were included regardless of drug role (suspect, concomitant, or interacting). The following agents were included:

1. Tyrosine kinase inhibitors (TKIs): Erlotinib, gefitinib, afatinib, osimertinib, dacomitinib, mobocertinib
2. Monoclonal antibodies / Other Biologics: Cetuximab, panitumumab, necitumumab, amivantamab, depatuxizumab mafodotin
3. Multi-kinase inhibitor with EGFR Activity: Vandetanib was included due to its documented secondary EGFR inhibition; however, its primary activity against vascular endothelial growth factor (VEGF) is acknowledged as a potential source of confounding [26].

For each drug, the total number of FAERS reports mentioning the drug (for any adverse event) was obtained.

### 2.3 Outcome Definition: Ocular Adverse Events

A list of clinician-relevant ocular adverse events (AEs) was also pre-specified as targets with which FAERS was queried. This list was then mapped to the corresponding Medical Dictionary for Regulatory Activities (MedDRA) Preferred Terms (PTs) used to query the `[patient.reaction.reactionmeddrapt]` field. Two AEs were consolidated to account for terminology differences. ‘‘DRY EYE SYNDROME’ and ‘DRY EYE DISEASE’ were mapped to ‘DRY EYE’. Additionally, ‘ACQUIRED TRICHOMEGALY’ was included with ‘TRICHOMEGALY’. The final, comprehensive PT list used is available in the **Supplementary Material (Table S1)**.

### 2.4 Disproportionality Analysis

For each drug-event pair, we constructed a 2×2 contingency table from FAERS counts:

- a: reports with both the drug and the event
- b: reports with the drug but not the event
- c: reports with the event but not the drug
- d: all other reports

We then calculated the Proportional Reporting Ratio (PRR) for each combination:

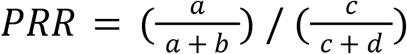

The PRR is a standard measure of disproportionality used in pharmacovigilance to identify potential safety signals [25, 27].

This compares the proportion of a specific event among reports for a given drug to the proportion of that event among reports for all other drugs. Two-sided p values were calculated using chi-square tests with Fisher’s exact test when any cell count was <5, and 95% confidence intervals (CIs) for PRR were derived. Because the variance of reporting ratios is high for drug, event combinations with small numbers of cases and to account for multiple comparisons, we applied the Benjamini-Hochberg false discovery rate (FDR) correction [28].

### 2.5 Signal Detection Criteria

A potential safety signal was defined based on conventional criteria used in FDA pharmacovigilance [25, 27]:

1. A minimum of ≥3 co-reported cases (a ≥3)
2. A PRR ≥ 2.0, indicating the event was reported at least twice as frequently with the drug compared to other drugs
3. A Benjamini-Hochberg FDR-adjusted p-value (q-value) <0.05

Drug/Event pairs meeting all three criteria were considered significant signals and formed the basis for subsequent clinical interpretation.

Drug/Events combinations

## Results

The pharmacovigilance query of FAERS database analyzed a total of 6,976,462 drug/event combinations. We identified potential safety signals for ocular adverse events (AEs) associated with 12 EGFR-targeted anticancer agents. Of the combinations examined, 20 pairs met all three signal detection criteria (a ≥3, PRR ≥2.0, FDR p<0.05). The strength and distribution of these significant signals are summarized in **Figure 1** and visualized in **Figure 2**. The full dataset of all analyzed drug/event combinations is provided in the **Supplementary Material (Table S2)**.

**Figure 1.**
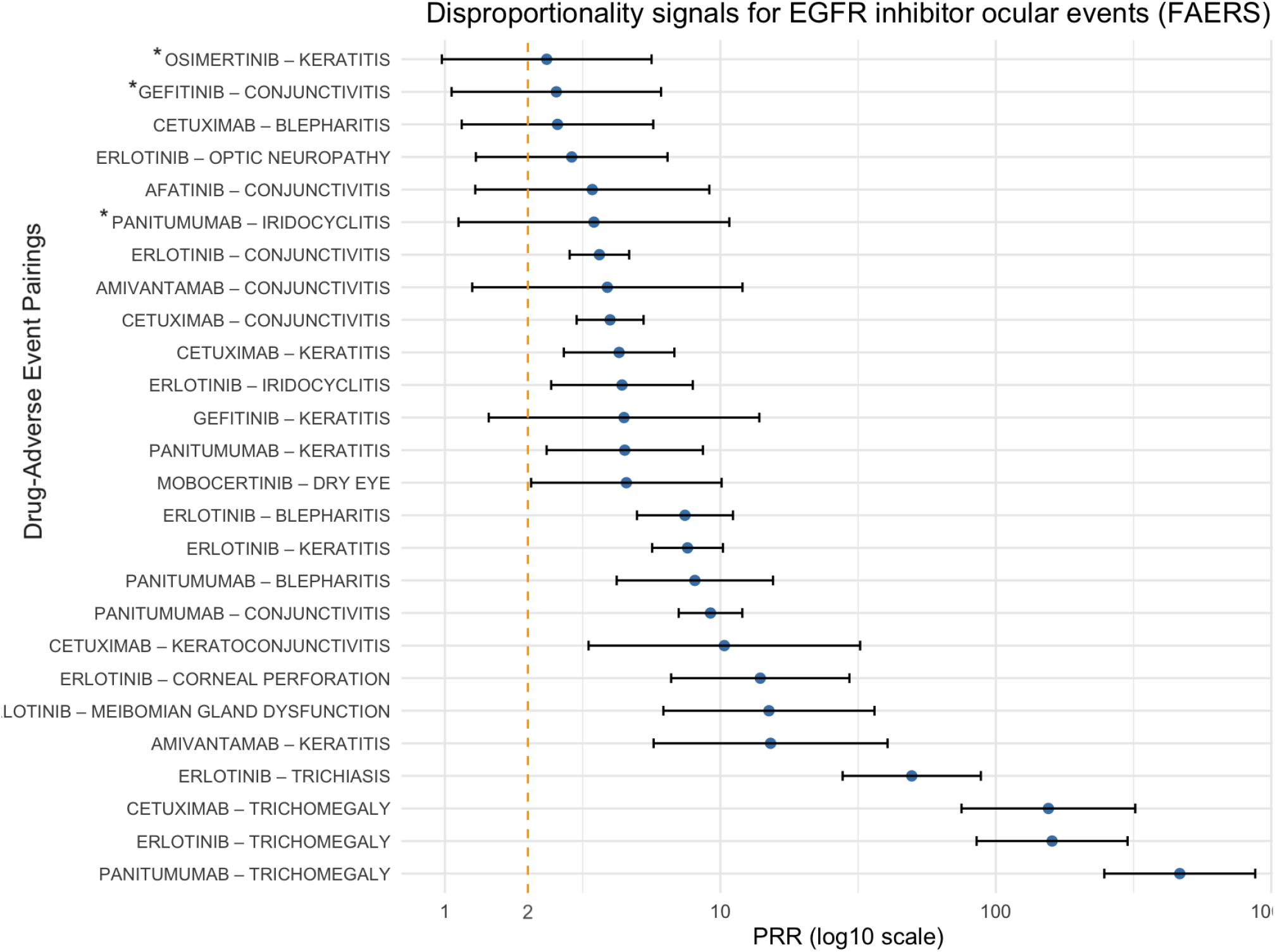
Summative Forest Plot of EGFR Agents with relevant Aes The dashed line represents the PRR significance threshold (2). Osimertinib-keratitis, gefitinib-conjunctivitis, and panitumumab-iridocyclitis, met the threshold for PRR and co-reported but are considered insufficient signals as their q-FDR levels were beyond the strict cut-off (indicated by the asterisk*).

**Figure 2.**
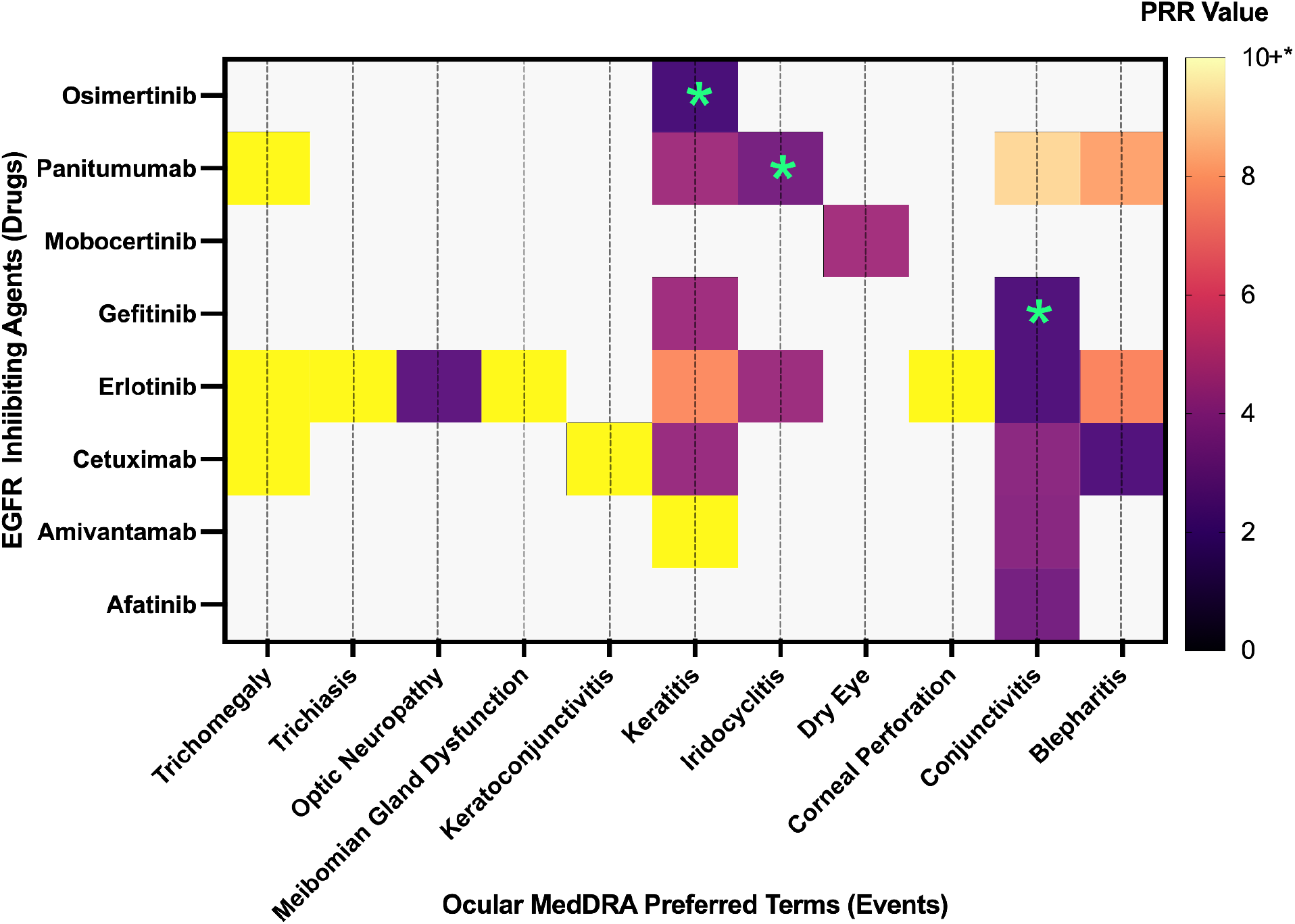
Heatmap Representation of Ocular Adverse Event Disproportionality. Note that the clear spaces indicate no events and that the (asterisk*) overlying three locations indicates the only situations in which the FDR threshold for a signal was *not* met despite having ≥2 PRR and ≥3 events.

### Drug-Specific Signal Patterns

Distinct patterns of ocular toxicity emerged among the agents analyzed.

- Panitumumab: Exhibited a dual toxicity profile characterized by severe eyelash abnormalities (highest trichomegaly PRR) and extensive ocular surface inflammation
- Cetuximab: Similarly showed strong signals for trichomegaly and surface disease, particularly blepharitis and conjunctivitis
- Erlotinib: Demonstrated ocular poly-toxicity, with significant signals spanning the entire spectrum from eyelid changes to vision-threatening corneal perforation and optic neuropathy
- Amivantamab: As a novel bispecific antibody, it displayed a highly specific and intense signal for keratitis (PRR= 15.2, 95% CI= 5.7-40.5) compared to other agents.
- Osimertinib: Showed a comparatively safer ocular profile; while a signal for keratitis was detected, it did not meet the strict FDR significance threshold in this analysis (PRR= 2.3, n= 5)
- Mobocertinib: Was uniquely associated with a specific signal for dry eye disease
- Dacomitinib, depatuxizumab, necitumumab, & vandetanib drug-event crossovers were largely absent, not reaching the significance thresholds.

More broadly, the signals clustered into three distinct clinical domains: (i) eyelash abnormalities, (ii) ocular surface disease, and (iii) vision-threatening intraocular pathology.

### i. Eyelash Abnormalities

The most pronounced disproportionality was observed for eyelash disorders. Trichomegaly displayed exceptionally high reporting associations across the FAERS database, with 89 reports; of these, 30 (33.7%) involved EGFR inhibitors. The PRR values were markedly elevated for panitumumab (PRR= 465.3, 95% CI= 247.7-874.3), erlotinib (PRR= 160.0, 95% CI= 85.1-300.8), and cetuximab (PRR= 155.2, 95% CI= 75.1-320.8). A significant signal for trichiasis was also identified, primarily associated with erlotinib (12 cases; PRR= 49.5, 95% CI= 27.8-88.2).

### ii. Ocular Surface Disease

A consistent pattern of ocular surface toxicity emerged across multiple agents, suggesting a class-wide effect. Conjunctivitis and keratitis were the most frequently reported events. As detailed in **Table 1**, strong signals for conjunctivitis were found for panitumumab, cetuximab, and erlotinib. Similarly, keratitis signals were significant for several agents, with the highest PRR observed for amivantamab, followed by erlotinib. Other ocular surface events exhibiting disproportionate reporting included dry eye (mobocertinib) and meibomian gland dysfunction (erlotinib).

**Table 1.** Ocular Surface Disease Signals. Grouped by Event and Sorted by Descending PRR. Showing only those that met the case count, PRR value and FDR criterion thresholds. CI= 95% Confidence Interval.

| Event (MedDRA PT) | Drug | Cases (n) | PRR | Lower CI | Upper CI | q-Value |
| --- | --- | --- | --- | --- | --- | --- |
| Conjunctivitis |  |  |  |  |  |  |
|  | Panitumumab | 54 | 9.21 | 7.06 | 12.01 | <.001 |
|  | Cetuximab | 49 | 3.98 | 3.01 | 5.26 | <.001 |
|  | Erlotinib | 62 | 3.64 | 2.84 | 4.66 | <.001 |
|  | Amivantamab | 3 | 3.89 | 1.26 | 12.03 | 0.04 |
|  | Afatinib | 4 | 3.43 | 1.29 | 9.12 | 0.04 |
| Keratitis |  |  |  |  |  |  |
|  | Amivantamab | 4 | 15.2 | 5.72 | 40.45 | <.001 |
|  | Erlotinib | 44 | 7.60 | 5.65 | 10.22 | <.001 |
|  | Gefitinib | 3 | 4.47 | 1.44 | 13.85 | 0.04 |
|  | Panitumumab | 9 | 4.50 | 2.34 | 8.64 | <.001 |
|  | Cetuximab | 18 | 4.29 | 2.70 | 6.81 | <.001 |
| Blepharitis |  |  |  |  |  |  |
|  | Panitumumab | 9 | 8.08 | 4.20 | 15.53 | <.001 |
|  | Erlotinib | 24 | 7.44 | 4.98 | 11.11 | <.001 |
|  | Cetuximab | 6 | 2.56 | 1.15 | 5.71 | 0.05 |
| Other |  |  |  |  |  |  |
| Meibomian Gland Dysfunction | Erlotinib | 5 | 15.00 | 6.21 | 36.26 | <.001 |
| Dry Eye | Mobocertinib | 6 | 4.56 | 2.06 | 10.10 | .004 |

### iii. Vision-Threatening and Intraocular Events

Less frequent but clinically serious events also demonstrated significant disproportionality. Corneal perforation was associated with erlotinib (7 cases; PRR= 13.9, 95% CI= 6.6-29.4). A signal for iridocyclitis was detected for erlotinib (11 cases; PRR= 4.3, 95% CI= 2.4-7.9). Additionally, a signal for optic neuropathy was identified with erlotinib (6 cases; PRR= 2.9, 95% CI= 1.3-6.4).

Of note, three drug-event pairs met the initial thresholds for case count (a ≥3) and PRR (≥2.0) but were not statistically significant after FDR adjustment (q-value ≥0.05): gefitinib with conjunctivitis (PRR= 2.5), panitumumab with iridocyclitis (PRR= 3.5), and osimertinib with keratitis (PRR= 2.3). These are noted in **Figure 1** and were not included in the primary signal count.

## Discussion

This comprehensive disproportionality analysis of the FAERS database identifies a diverse spectrum of ocular safety signals associated with EGFR-targeted therapies. The most pronounced signals were observed for eyelash abnormalities, particularly trichomegaly, with exceptionally high PRR values for panitumumab, erlotinib, and cetuximab. A consistent pattern of ocular surface inflammation, including conjunctivitis, keratitis, and blepharitis, was evident across multiple agents, suggesting a class-wide effect. Importantly, significant signals were detected for serious, vision-threatening events such as corneal perforation, iridocyclitis, and optic neuropathy, primarily associated with erlotinib. To our knowledge, this is the first FDR-controlled, class-wide FAERS disproportionality analysis of EGFR inhibitors that includes newer biologics such as amivantamab and necitumumab, and it newly highlights keratitis and dry eye signals for agents such as amivantamab and mobocertinib that clinicians may not routinely associate with ocular toxicity. However, as exposures increase for the newer EGFR biologics, report counts and signal stability are expected to increase, though early disproportionality may also be impacted by increased initial reporting.

These findings corroborate and extend the evolving understanding of EGFR inhibitor ocular toxicity. The data reveal that EGFR inhibitors are responsible for a substantial proportion of specific drug-induced ocular events; notably, of the 89 total reports of trichomegaly in the FAERS database, 30 (33.7%) were attributed to EGFR inhibitors. The strong signals for trichomegaly and trichiasis quantitatively confirm the extensive case report literature [7, 10, 18, 19], while the signals for rare intraocular events highlight a potential risk profile that has been less comprehensively documented.

The biological plausibility of these findings is sound and supports the validity of the detected signals. Specifically, EGFR is constitutively expressed in the basal epithelial cells of the cornea, conjunctiva, meibomian glands, and hair follicles [4, 13, 17]. Given EGFR’s role in cellular proliferation, impaired corneal epithelial healing leading to keratitis and potential perforation help explains the observed toxicity. The dysfunction of the meibomian glands contributes to dry eye and blepharitis and dysregulation of the eyelash growth cycle resulting in trichomegaly [11, 14, 17, 18]. The concentration of these events across both monoclonal antibodies and small-molecule TKIs reinforces that the common pathway is EGFR blockade itself.

It is crucial to emphasize that the PRR signals quantified here indicate a disproportionate reporting association, not a proven causal risk. As outlined in the FDA’s framework for data mining [25] and recent methodological reviews [29], disproportionality analysis is a hypothesis-generating tool. The statistical rigor of our study was enhanced by applying a false discovery rate (FDR) correction to mitigate the risk of false positives inherent in multiple comparisons, a step that adds confidence to the signals we report [28]. Still, these findings must be interpreted within the inherent constraints of spontaneous reporting systems.

The clinical implications of these signals are significant. Given the class-wide nature of ocular surface disease and the potential for serious corneal complications, a proactive approach to ocular monitoring is warranted. We recommend that patients initiating EGFR inhibitor therapy undergo a baseline ophthalmologic assessment; this practice, suggested in prior reviews [22, 23], can establish a reference point, educate patients on early symptom recognition, and anchor subsequent follow-up. For example, patients should be counseled to report new ocular surface discomfort promptly, and clinicians should consider low-threshold referral to ophthalmology for persistent keratitis, trichomegaly causing lash-cornea touch, or visual symptoms. Regular monitoring throughout treatment, especially with newer, potentially higher-risk agents, may facilitate early intervention, thereby preserving visual function and quality of life and preventing interruptions in life-saving cancer therapy.

### Limitations & Ethics

The interpretation of our results must be contextualized by the well-documented limitations of pharmacovigilance studies. FAERS is a passive surveillance system susceptible to under-reporting and variable data quality [25, 29], and event misclassification is possible; free-text narratives and MedDRA coding may under-capture relevant ocular AEs, particularly when they are grouped under nonspecific terms such as ‘eye disorder.’ Because we relied on the openFDA `[safetyreportid]` without further manual de-duplication, residual duplicate reports, common when a case is updated by different sources, may exist. While this might inflate absolute case counts, it is unlikely to materially alter the qualitative pattern of signals detected.

Disproportionality analysis also lacks a true denominator (i.e., the total number of patients exposed) and information on time-to-onset or dechallenge, precluding incidence estimates and limiting inferences about temporal relationships; the PRR therefore reflects reporting association, not absolute risk. In addition, we included all drug roles (suspect, concomitant, and interacting), which may dilute specificity of the signals but minimizes the chance of missing relevant reports.

Furthermore, our analysis compared EGFR inhibitors against the full background of all drugs in FAERS. While standard, this method can be susceptible to comparator bias; however, the magnitude of the signals observed here suggests they are unlikely to be artifacts of the background rate. The ‘Weber effect,’ whereby reporting rates are elevated for newly marketed drugs due to increased vigilance, may partially influence the signals for newer agents [30, 31]. However, the signal for amivantamab-associated keratitis (PRR= 15.2) is of a magnitude that exceeds typical Weber-effect biases and still warrants cautious interpretation rather than dismissal as pure reporting artifact.

Finally, confounding remains a critical challenge in oncology [32]. Patients often receive complex multi-drug regimens and have systemic comorbidities; attributing an event such as optic neuropathy solely to an EGFR inhibitor is difficult given the potential neurotoxicity of concurrent platinum-based chemotherapy or cranial irradiation [33], and uveitis and orbital inflammation are also associated with immune checkpoint inhibitors [34]. Therefore, while these signals warrant vigilance, they do not definitively establish causality.

Because FAERS data are publicly available and fully de-identified, this analysis did not require institutional review board approval under 45 CFR § 46.104. This study was conducted in accordance with the 1964 Helsinki Declaration and its ethical standards.

## Conclusion

This large-scale pharmacovigilance study provides robust, updated evidence of a class-wide spectrum of ocular toxicity associated with EGFR inhibitors. While these findings represent disproportionality signals rather than incidence rates, the consistent patterns observed, ranging from eyelid changes to sight-threatening corneal events, are clinically significant. These results support the implementation of proactive ocular monitoring and heightened vigilance, particularly for patients on newer agents like amivantamab. Future efforts should prioritize ophthalmology-oncology collaboration and prospective studies to determine true incidence rates and standardize management guidelines.

## Supporting information

Combined Supplementary Material

## Data Availability

The US Food and Drug Administration (FDA) Adverse Event Reporting System (FAERS) is publicly available through openFDA.

https://open.fda.gov/

## Acknowledgements

The author(s) have no financial disclosures or conflicts of interest to report for this study.

