## Supplementary material for "Evolving Ocular Safety Signals of EGFR Inhibitors: A FAERS Disproportionality Analysis of Amivantamab, Mobocertinib, and Classic Agents": Combined Supplementary Material

**Table S1.**
List of the Adverse Event search terms used within the database.

| Search Terms Used | MedDRA Grouping |
|---|---|
| DRY EYE SYNDROME | MEDDRA/10013777 |
| DRY EYE DISEASE | MEDDRA/10013774 |
| MEIBOMIAN GLAND DYSFUNCTION | MEDDRA/10065062 |
| MEIBOMITIS | MEDDRA/10052157 |
| BLEPHARITIS | MEDDRA/10005148 |
| CONJUNCTIVITIS | MEDDRA/10010741 |
| KERATOCONJUNCTIVITIS | MEDDRA/10023348 |
| KERATITIS | MEDDRA/10023332 |
| CORNEAL ULCER | MEDDRA/10048492 |
| CORNEAL EROSION | MEDDRA/10011013 |
| CORNEAL ULCERATION | MEDDRA/10011060 |
| CORNEAL EPITHELIAL DEFECT | **MEDDRA/10075399** |
| PERSISTENT CORNEAL EPITHELIAL DEFECT | **MEDDRA/10075399** |
| CORNEAL MELT | MEDDRA/10059795 |
| CORNEAL PERFORATION | MEDDRA/10011039 |
| ACQUIRED TRICHOMEGALY | **MEDDRA/10044613** |
| “TRICHOMEGALY” | **MEDDRA/10044613** |
| TRICHIASIS | MEDDRA/10044604 |
| LID ECTROPION | MEDDRA/10014179 |
| IRIDOCYCLITIS | MEDDRA/10022941 |
| UVEITIS | MEDDRA/10046851 |
| EYE PAIN | MEDDRA/10015958 |
| PHOTOPHOBIA | MEDDRA/10034960 |
| SUBCONJUNCTIVAL HEMORRHAGE | MEDDRA/10055345 |
| CONJUNCTIVAL HEMORRHAGE | MEDDRA/10010720 |
| RETINAL HEMORRHAGE | MEDDRA/10038870 |
| OPTIC NEURITIS | MEDDRA/10030942 |
| OPTIC NEUROPATHY | MEDDRA/10061323 |
| VISUAL ACUITY REDUCED | MEDDRA/10047531 |
| VISION LOSS | MEDDRA/10047522 |
| VORTEX KERATOPATHY | MEDDRA/10077621 |
| CORNEAL VERTICILLATA | MEDDRA/10077621 |

| **Table S2.** Complete Signal Data | | | | | | | | | | |
|---|---|---|---|---|---|---|---|---|---|---|
| Drug | Event | Label | total_drug_reports | total_event_reports | a | total_faers | prr | b | c | d |
| ERLOTINIB | BLEPHARITIS | Erlotinib / Tarceva | 17333 | 3685 | 24 | 19684585 | 7.438433047 | 17309 | 3661 | 19663591 |
| ERLOTINIB | CONJUNCTIVAL HEMORRHAGE | Erlotinib / Tarceva | 17333 | -Inf | NA | 19684585 | NA | NA | NA | NA |
| ERLOTINIB | CONJUNCTIVITIS | Erlotinib / Tarceva | 17333 | 19405 | 62 | 19684585 | 3.636953958 | 17271 | 19343 | 19647909 |
| ERLOTINIB | CORNEAL EPITHELIAL DEFECT | Erlotinib / Tarceva | 17333 | 99 | 1 | 19684585 | 11.57827525 | 17332 | 98 | 19667154 |
| ERLOTINIB | CORNEAL EROSION | Erlotinib / Tarceva | 17333 | 496 | 2 | 19684585 | 4.593809613 | 17331 | 494 | 19666758 |
| ERLOTINIB | CORNEAL MELT | Erlotinib / Tarceva | 17333 | -Inf | NA | 19684585 | NA | NA | NA | NA |
| ERLOTINIB | CORNEAL PERFORATION | Erlotinib / Tarceva | 17333 | 576 | 7 | 19684585 | 13.95904538 | 17326 | 569 | 19666683 |
| ERLOTINIB | CORNEAL ULCER | Erlotinib / Tarceva | 17333 | 101 | NA | 19684585 | NA | NA | NA | NA |
| ERLOTINIB | CORNEAL ULCERATION | Erlotinib / Tarceva | 17333 | -Inf | NA | 19684585 | NA | NA | NA | NA |
| ERLOTINIB | CORNEAL VERTICILLATA | Erlotinib / Tarceva | 17333 | -Inf | NA | 19684585 | NA | NA | NA | NA |
| ERLOTINIB | DRY EYE | Erlotinib / Tarceva | 17333 | 43203 | 62 | 19684585 | 1.630690072 | 17271 | 43141 | 19624111 |
| ERLOTINIB | EYE PAIN | Erlotinib / Tarceva | 17333 | 51035 | 26 | 19684585 | 0.5783576493 | 17307 | 51009 | 19616243 |
| ERLOTINIB | IRIDOCYCLITIS | Erlotinib / Tarceva | 17333 | 2852 | 11 | 19684585 | 4.393305427 | 17322 | 2841 | 19664411 |
| ERLOTINIB | KERATITIS | Erlotinib / Tarceva | 17333 | 6613 | 44 | 19684585 | 7.600170936 | 17289 | 6569 | 19660683 |
| ERLOTINIB | KERATOCONJUNCTIVITIS | Erlotinib / Tarceva | 17333 | 459 | NA | 19684585 | NA | NA | NA | NA |
| ERLOTINIB | LID ECTROPION | Erlotinib / Tarceva | 17333 | -Inf | NA | 19684585 | NA | NA | NA | NA |
| ERLOTINIB | MEIBOMIAN GLAND DYSFUNCTION | Erlotinib / Tarceva | 17333 | 383 | 5 | 19684585 | 15.00887532 | 17328 | 378 | 19666874 |
| ERLOTINIB | OPTIC NEURITIS | Erlotinib / Tarceva | 17333 | 8787 | 4 | 19684585 | 0.516757816 | 17329 | 8783 | 19658469 |
| ERLOTINIB | OPTIC NEUROPATHY | Erlotinib / Tarceva | 17333 | 2365 | 6 | 19684585 | 2.885979587 | 17327 | 2359 | 19664893 |
| ERLOTINIB | PERSISTENT CORNEAL EPITHELIAL DEFECT | Erlotinib / Tarceva | 17333 | 99 | 1 | 19684585 | 11.57827525 | 17332 | 98 | 19667154 |
| ERLOTINIB | PHOTOPHOBIA | Erlotinib / Tarceva | 17333 | 17555 | 8 | 19684585 | 0.5173173645 | 17325 | 17547 | 19649705 |
| ERLOTINIB | RETINAL HEMORRHAGE | Erlotinib / Tarceva | 17333 | -Inf | NA | 19684585 | NA | NA | NA | NA |
| ERLOTINIB | SUBCONJUNCTIVAL HEMORRHAGE | Erlotinib / Tarceva | 17333 | -Inf | NA | 19684585 | NA | NA | NA | NA |
| ERLOTINIB | TRICHIASIS | Erlotinib / Tarceva | 17333 | 287 | 12 | 19684585 | 49.51291525 | 17321 | 275 | 19666977 |
| ERLOTINIB | TRICHOMEGALY | Erlotinib / Tarceva | 17333 | 89 | 11 | 19684585 | 160.0177015 | 17322 | 78 | 19667174 |
| ERLOTINIB | UVEITIS | Erlotinib / Tarceva | 17333 | 13175 | 11 | 19684585 | 0.9481449954 | 17322 | 13164 | 19654088 |
| ERLOTINIB | VISION LOSS | Erlotinib / Tarceva | 17333 | 193 | NA | 19684585 | NA | NA | NA | NA |
| ERLOTINIB | VISUAL ACUITY REDUCED | Erlotinib / Tarceva | 17333 | 32697 | 34 | 19684585 | 1.18111665 | 17299 | 32663 | 19634589 |
| ERLOTINIB | VORTEX KERATOPATHY | Erlotinib / Tarceva | 17333 | -Inf | NA | 19684585 | NA | NA | NA | NA |
| GEFITINIB | BLEPHARITIS | Gefitinib / Iressa | 1999 | 3685 | NA | 19684585 | NA | NA | NA | NA |
| GEFITINIB | CONJUNCTIVAL HEMORRHAGE | Gefitinib / Iressa | 1999 | -Inf | NA | 19684585 | NA | NA | NA | NA |
| GEFITINIB | CONJUNCTIVITIS | Gefitinib / Iressa | 1999 | 19405 | 5 | 19684585 | 2.537684564 | 1994 | 19400 | 19663186 |
| GEFITINIB | CORNEAL EPITHELIAL DEFECT | Gefitinib / Iressa | 1999 | 99 | NA | 19684585 | NA | NA | NA | NA |
| GEFITINIB | CORNEAL EROSION | Gefitinib / Iressa | 1999 | 496 | NA | 19684585 | NA | NA | NA | NA |
| GEFITINIB | CORNEAL MELT | Gefitinib / Iressa | 1999 | -Inf | NA | 19684585 | NA | NA | NA | NA |
| GEFITINIB | CORNEAL PERFORATION | Gefitinib / Iressa | 1999 | 576 | 1 | 19684585 | 17.1238541 | 1998 | 575 | 19682011 |
| GEFITINIB | CORNEAL ULCER | Gefitinib / Iressa | 1999 | 101 | NA | 19684585 | NA | NA | NA | NA |
| GEFITINIB | CORNEAL ULCERATION | Gefitinib / Iressa | 1999 | -Inf | NA | 19684585 | NA | NA | NA | NA |
| GEFITINIB | CORNEAL VERTICILLATA | Gefitinib / Iressa | 1999 | -Inf | NA | 19684585 | NA | NA | NA | NA |
| GEFITINIB | DRY EYE | Gefitinib / Iressa | 1999 | 43203 | 3 | 19684585 | 0.6837650075 | 1996 | 43200 | 19639386 |
| GEFITINIB | EYE PAIN | Gefitinib / Iressa | 1999 | 51035 | NA | 19684585 | NA | NA | NA | NA |
| GEFITINIB | IRIDOCYCLITIS | Gefitinib / Iressa | 1999 | 2852 | NA | 19684585 | NA | NA | NA | NA |
| GEFITINIB | KERATITIS | Gefitinib / Iressa | 1999 | 6613 | 3 | 19684585 | 4.468781895 | 1996 | 6610 | 19675976 |
| GEFITINIB | KERATOCONJUNCTIVITIS | Gefitinib / Iressa | 1999 | 459 | NA | 19684585 | NA | NA | NA | NA |
| GEFITINIB | LID ECTROPION | Gefitinib / Iressa | 1999 | -Inf | NA | 19684585 | NA | NA | NA | NA |
| GEFITINIB | MEIBOMIAN GLAND DYSFUNCTION | Gefitinib / Iressa | 1999 | 383 | NA | 19684585 | NA | NA | NA | NA |
| GEFITINIB | OPTIC NEURITIS | Gefitinib / Iressa | 1999 | 8787 | NA | 19684585 | NA | NA | NA | NA |
| GEFITINIB | OPTIC NEUROPATHY | Gefitinib / Iressa | 1999 | 2365 | NA | 19684585 | NA | NA | NA | NA |
| GEFITINIB | PERSISTENT CORNEAL EPITHELIAL DEFECT | Gefitinib / Iressa | 1999 | 99 | NA | 19684585 | NA | NA | NA | NA |
| GEFITINIB | PHOTOPHOBIA | Gefitinib / Iressa | 1999 | 17555 | 1 | 19684585 | 0.5609101121 | 1998 | 17554 | 19665032 |
| GEFITINIB | RETINAL HEMORRHAGE | Gefitinib / Iressa | 1999 | -Inf | NA | 19684585 | NA | NA | NA | NA |
| GEFITINIB | SUBCONJUNCTIVAL HEMORRHAGE | Gefitinib / Iressa | 1999 | -Inf | NA | 19684585 | NA | NA | NA | NA |

| **Table S2.** Complete Signal Data | | | | | | | | | | |
|---|---|---|---|---|---|---|---|---|---|---|
| Drug | Event | Label | total_drug_reports | total_event_reports | a | total_faers | prr | b | c | d |
| GEFITINIB | TRICHIASIS | Gefitinib / Iressa | 1999 | 287 | NA | 19684585 | NA | NA | NA | NA |
| GEFITINIB | TRICHOMEGALY | Gefitinib / Iressa | 1999 | 89 | NA | 19684585 | NA | NA | NA | NA |
| GEFITINIB | UVEITIS | Gefitinib / Iressa | 1999 | 13175 | 1 | 19684585 | 0.7473976095 | 1998 | 13174 | 19669412 |
| GEFITINIB | VISION LOSS | Gefitinib / Iressa | 1999 | 193 | NA | 19684585 | NA | NA | NA | NA |
| GEFITINIB | VISUAL ACUITY REDUCED | Gefitinib / Iressa | 1999 | 32697 | NA | 19684585 | NA | NA | NA | NA |
| GEFITINIB | VORTEX KERATOPATHY | Gefitinib / Iressa | 1999 | -Inf | NA | 19684585 | NA | NA | NA | NA |
| AFATINIB | BLEPHARITIS | Afatinib / Gilotrif | 1184 | 3685 | NA | 19684585 | NA | NA | NA | NA |
| AFATINIB | CONJUNCTIVAL HEMORRHAGE | Afatinib / Gilotrif | 1184 | -Inf | NA | 19684585 | NA | NA | NA | NA |
| AFATINIB | CONJUNCTIVITIS | Afatinib / Gilotrif | 1184 | 19405 | 4 | 19684585 | 3.427554062 | 1180 | 19401 | 19664000 |
| AFATINIB | CORNEAL EPITHELIAL DEFECT | Afatinib / Gilotrif | 1184 | 99 | NA | 19684585 | NA | NA | NA | NA |
| AFATINIB | CORNEAL EROSION | Afatinib / Gilotrif | 1184 | 496 | NA | 19684585 | NA | NA | NA | NA |
| AFATINIB | CORNEAL MELT | Afatinib / Gilotrif | 1184 | -Inf | NA | 19684585 | NA | NA | NA | NA |
| AFATINIB | CORNEAL PERFORATION | Afatinib / Gilotrif | 1184 | 576 | NA | 19684585 | NA | NA | NA | NA |
| AFATINIB | CORNEAL ULCER | Afatinib / Gilotrif | 1184 | 101 | NA | 19684585 | NA | NA | NA | NA |
| AFATINIB | CORNEAL ULCERATION | Afatinib / Gilotrif | 1184 | -Inf | NA | 19684585 | NA | NA | NA | NA |
| AFATINIB | CORNEAL VERTICILLATA | Afatinib / Gilotrif | 1184 | -Inf | NA | 19684585 | NA | NA | NA | NA |
| AFATINIB | DRY EYE | Afatinib / Gilotrif | 1184 | 43203 | 2 | 19684585 | 0.7696346884 | 1182 | 43201 | 19640200 |
| AFATINIB | EYE PAIN | Afatinib / Gilotrif | 1184 | 51035 | 2 | 19684585 | 0.6515193733 | 1182 | 51033 | 19632368 |
| AFATINIB | IRIDOCYCLITIS | Afatinib / Gilotrif | 1184 | 2852 | NA | 19684585 | NA | NA | NA | NA |
| AFATINIB | KERATITIS | Afatinib / Gilotrif | 1184 | 6613 | 2 | 19684585 | 5.029343242 | 1182 | 6611 | 19676790 |
| AFATINIB | KERATOCONJUNCTIVITIS | Afatinib / Gilotrif | 1184 | 459 | NA | 19684585 | NA | NA | NA | NA |
| AFATINIB | LID ECTROPION | Afatinib / Gilotrif | 1184 | -Inf | NA | 19684585 | NA | NA | NA | NA |
| AFATINIB | MEIBOMIAN GLAND DYSFUNCTION | Afatinib / Gilotrif | 1184 | 383 | NA | 19684585 | NA | NA | NA | NA |
| AFATINIB | OPTIC NEURITIS | Afatinib / Gilotrif | 1184 | 8787 | NA | 19684585 | NA | NA | NA | NA |
| AFATINIB | OPTIC NEUROPATHY | Afatinib / Gilotrif | 1184 | 2365 | NA | 19684585 | NA | NA | NA | NA |
| AFATINIB | PERSISTENT CORNEAL EPITHELIAL DEFECT | Afatinib / Gilotrif | 1184 | 99 | NA | 19684585 | NA | NA | NA | NA |
| AFATINIB | PHOTOPHOBIA | Afatinib / Gilotrif | 1184 | 17555 | NA | 19684585 | NA | NA | NA | NA |
| AFATINIB | RETINAL HEMORRHAGE | Afatinib / Gilotrif | 1184 | -Inf | NA | 19684585 | NA | NA | NA | NA |
| AFATINIB | SUBCONJUNCTIVAL HEMORRHAGE | Afatinib / Gilotrif | 1184 | -Inf | NA | 19684585 | NA | NA | NA | NA |
| AFATINIB | TRICHIASIS | Afatinib / Gilotrif | 1184 | 287 | NA | 19684585 | NA | NA | NA | NA |
| AFATINIB | TRICHOMEGALY | Afatinib / Gilotrif | 1184 | 89 | NA | 19684585 | NA | NA | NA | NA |
| AFATINIB | UVEITIS | Afatinib / Gilotrif | 1184 | 13175 | 1 | 19684585 | 1.261916964 | 1183 | 13174 | 19670227 |
| AFATINIB | VISION LOSS | Afatinib / Gilotrif | 1184 | 193 | NA | 19684585 | NA | NA | NA | NA |
| AFATINIB | VISUAL ACUITY REDUCED | Afatinib / Gilotrif | 1184 | 32697 | 1 | 19684585 | 0.5084565111 | 1183 | 32696 | 19650705 |
| AFATINIB | VORTEX KERATOPATHY | Afatinib / Gilotrif | 1184 | -Inf | NA | 19684585 | NA | NA | NA | NA |
| OSIMERTINIB | BLEPHARITIS | Osimertinib / Tagrisso | 6359 | 3685 | 1 | 19684585 | 0.8399965407 | 6358 | 3684 | 19674542 |
| OSIMERTINIB | CONJUNCTIVAL HEMORRHAGE | Osimertinib / Tagrisso | 6359 | -Inf | NA | 19684585 | NA | NA | NA | NA |
| OSIMERTINIB | CONJUNCTIVITIS | Osimertinib / Tagrisso | 6359 | 19405 | 7 | 19684585 | 1.11670434 | 6352 | 19398 | 19658828 |
| OSIMERTINIB | CORNEAL EPITHELIAL DEFECT | Osimertinib / Tagrisso | 6359 | 99 | NA | 19684585 | NA | NA | NA | NA |
| OSIMERTINIB | CORNEAL EROSION | Osimertinib / Tagrisso | 6359 | 496 | NA | 19684585 | NA | NA | NA | NA |
| OSIMERTINIB | CORNEAL MELT | Osimertinib / Tagrisso | 6359 | -Inf | NA | 19684585 | NA | NA | NA | NA |
| OSIMERTINIB | CORNEAL PERFORATION | Osimertinib / Tagrisso | 6359 | 576 | NA | 19684585 | NA | NA | NA | NA |
| OSIMERTINIB | CORNEAL ULCER | Osimertinib / Tagrisso | 6359 | 101 | NA | 19684585 | NA | NA | NA | NA |
| OSIMERTINIB | CORNEAL ULCERATION | Osimertinib / Tagrisso | 6359 | -Inf | NA | 19684585 | NA | NA | NA | NA |
| OSIMERTINIB | CORNEAL VERTICILLATA | Osimertinib / Tagrisso | 6359 | -Inf | NA | 19684585 | NA | NA | NA | NA |
| OSIMERTINIB | DRY EYE | Osimertinib / Tagrisso | 6359 | 43203 | 11 | 19684585 | 0.7881093678 | 6348 | 43192 | 19635034 |
| OSIMERTINIB | EYE PAIN | Osimertinib / Tagrisso | 6359 | 51035 | 3 | 19684585 | 0.1819180469 | 6356 | 51032 | 19627194 |
| OSIMERTINIB | IRIDOCYCLITIS | Osimertinib / Tagrisso | 6359 | 2852 | 1 | 19684585 | 1.085425204 | 6358 | 2851 | 19675375 |
| OSIMERTINIB | KERATITIS | Osimertinib / Tagrisso | 6359 | 6613 | 5 | 19684585 | 2.341515781 | 6354 | 6608 | 19671618 |
| OSIMERTINIB | KERATOCONJUNCTIVITIS | Osimertinib / Tagrisso | 6359 | 459 | NA | 19684585 | NA | NA | NA | NA |
| OSIMERTINIB | LID ECTROPION | Osimertinib / Tagrisso | 6359 | -Inf | NA | 19684585 | NA | NA | NA | NA |
| OSIMERTINIB | MEIBOMIAN GLAND DYSFUNCTION | Osimertinib / Tagrisso | 6359 | 383 | NA | 19684585 | NA | NA | NA | NA |

**Table S2.** Complete Signal Data

| Drug | Event | Label | total_drug_reports | total_event_reports | a | total_faers | prr | b | c | d |
|---|---|---|---|---|---|---|---|---|---|---|
| OSIMERTINIB | OPTIC NEURITIS | Osimertinib / Tagrisso | 6359 | 8787 | NA | 19684585 | NA | NA | NA | NA |
| OSIMERTINIB | OPTIC NEUROPATHY | Osimertinib / Tagrisso | 6359 | 2365 | 1 | 19684585 | 1.309030142 | 6358 | 2364 | 19675862 |
| OSIMERTINIB | PERSISTENT CORNEAL EPITHELIAL DEFECT | Osimertinib / Tagrisso | 6359 | 99 | NA | 19684585 | NA | NA | NA | NA |
| OSIMERTINIB | PHOTOPHOBIA | Osimertinib / Tagrisso | 6359 | 17555 | NA | 19684585 | NA | NA | NA | NA |
| OSIMERTINIB | RETINAL HEMORRHAGE | Osimertinib / Tagrisso | 6359 | -Inf | NA | 19684585 | NA | NA | NA | NA |
| OSIMERTINIB | SUBCONJUNCTIVAL HEMORRHAGE | Osimertinib / Tagrisso | 6359 | -Inf | NA | 19684585 | NA | NA | NA | NA |
| OSIMERTINIB | TRICHIASIS | Osimertinib / Tagrisso | 6359 | 287 | NA | 19684585 | NA | NA | NA | NA |
| OSIMERTINIB | TRICHOMEGALY | Osimertinib / Tagrisso | 6359 | 89 | NA | 19684585 | NA | NA | NA | NA |
| OSIMERTINIB | UVEITIS | Osimertinib / Tagrisso | 6359 | 13175 | 6 | 19684585 | 1.409923573 | 6353 | 13169 | 19665057 |
| OSIMERTINIB | VISION LOSS | Osimertinib / Tagrisso | 6359 | 193 | NA | 19684585 | NA | NA | NA | NA |
| OSIMERTINIB | VISUAL ACUITY REDUCED | Osimertinib / Tagrisso | 6359 | 32697 | 1 | 19684585 | 0.09464605015 | 6358 | 32696 | 19645530 |
| OSIMERTINIB | VORTEX KERATOPATHY | Osimertinib / Tagrisso | 6359 | -Inf | NA | 19684585 | NA | NA | NA | NA |
| DACOMITINIB | BLEPHARITIS | Dacomitinib / Vizimpro | 456 | 3685 | NA | 19684585 | NA | NA | NA | NA |
| DACOMITINIB | CONJUNCTIVAL HEMORRHAGE | Dacomitinib / Vizimpro | 456 | -Inf | NA | 19684585 | NA | NA | NA | NA |
| DACOMITINIB | CONJUNCTIVITIS | Dacomitinib / Vizimpro | 456 | 19405 | 1 | 19684585 | 2.224641804 | 455 | 19404 | 19664725 |
| DACOMITINIB | CORNEAL EPITHELIAL DEFECT | Dacomitinib / Vizimpro | 456 | 99 | NA | 19684585 | NA | NA | NA | NA |
| DACOMITINIB | CORNEAL EROSION | Dacomitinib / Vizimpro | 456 | 496 | NA | 19684585 | NA | NA | NA | NA |
| DACOMITINIB | CORNEAL MELT | Dacomitinib / Vizimpro | 456 | -Inf | NA | 19684585 | NA | NA | NA | NA |
| DACOMITINIB | CORNEAL PERFORATION | Dacomitinib / Vizimpro | 456 | 576 | NA | 19684585 | NA | NA | NA | NA |
| DACOMITINIB | CORNEAL ULCER | Dacomitinib / Vizimpro | 456 | 101 | NA | 19684585 | NA | NA | NA | NA |
| DACOMITINIB | CORNEAL ULCERATION | Dacomitinib / Vizimpro | 456 | -Inf | NA | 19684585 | NA | NA | NA | NA |
| DACOMITINIB | CORNEAL VERTICILLATA | Dacomitinib / Vizimpro | 456 | -Inf | NA | 19684585 | NA | NA | NA | NA |
| DACOMITINIB | DRY EYE | Dacomitinib / Vizimpro | 456 | 43203 | 2 | 19684585 | 1.998423627 | 454 | 43201 | 19640928 |
| DACOMITINIB | EYE PAIN | Dacomitinib / Vizimpro | 456 | 51035 | NA | 19684585 | NA | NA | NA | NA |
| DACOMITINIB | IRIDOCYCLITIS | Dacomitinib / Vizimpro | 456 | 2852 | NA | 19684585 | NA | NA | NA | NA |
| DACOMITINIB | KERATITIS | Dacomitinib / Vizimpro | 456 | 6613 | NA | 19684585 | NA | NA | NA | NA |
| DACOMITINIB | KERATOCONJUNCTIVITIS | Dacomitinib / Vizimpro | 456 | 459 | NA | 19684585 | NA | NA | NA | NA |
| DACOMITINIB | LID ECTROPION | Dacomitinib / Vizimpro | 456 | -Inf | NA | 19684585 | NA | NA | NA | NA |
| DACOMITINIB | MEIBOMIAN GLAND DYSFUNCTION | Dacomitinib / Vizimpro | 456 | 383 | 1 | 19684585 | 113.0024858 | 455 | 382 | 19683747 |
| DACOMITINIB | OPTIC NEURITIS | Dacomitinib / Vizimpro | 456 | 8787 | NA | 19684585 | NA | NA | NA | NA |
| DACOMITINIB | OPTIC NEUROPATHY | Dacomitinib / Vizimpro | 456 | 2365 | NA | 19684585 | NA | NA | NA | NA |
| DACOMITINIB | PERSISTENT CORNEAL EPITHELIAL DEFECT | Dacomitinib / Vizimpro | 456 | 99 | NA | 19684585 | NA | NA | NA | NA |
| DACOMITINIB | PHOTOPHOBIA | Dacomitinib / Vizimpro | 456 | 17555 | NA | 19684585 | NA | NA | NA | NA |
| DACOMITINIB | RETINAL HEMORRHAGE | Dacomitinib / Vizimpro | 456 | -Inf | NA | 19684585 | NA | NA | NA | NA |
| DACOMITINIB | SUBCONJUNCTIVAL HEMORRHAGE | Dacomitinib / Vizimpro | 456 | -Inf | NA | 19684585 | NA | NA | NA | NA |
| DACOMITINIB | TRICHIASIS | Dacomitinib / Vizimpro | 456 | 287 | NA | 19684585 | NA | NA | NA | NA |
| DACOMITINIB | TRICHOMEGALY | Dacomitinib / Vizimpro | 456 | 89 | NA | 19684585 | NA | NA | NA | NA |
| DACOMITINIB | UVEITIS | Dacomitinib / Vizimpro | 456 | 13175 | NA | 19684585 | NA | NA | NA | NA |
| DACOMITINIB | VISION LOSS | Dacomitinib / Vizimpro | 456 | 193 | NA | 19684585 | NA | NA | NA | NA |
| DACOMITINIB | VISUAL ACUITY REDUCED | Dacomitinib / Vizimpro | 456 | 32697 | 1 | 19684585 | 1.320251699 | 455 | 32696 | 19651433 |
| DACOMITINIB | VORTEX KERATOPATHY | Dacomitinib / Vizimpro | 456 | -Inf | NA | 19684585 | NA | NA | NA | NA |
| MOBOCERTINIB | BLEPHARITIS | Mobocertinib / EXKIVITY | 600 | 3685 | NA | 19684585 | NA | NA | NA | NA |
| MOBOCERTINIB | CONJUNCTIVAL HEMORRHAGE | Mobocertinib / EXKIVITY | 600 | -Inf | NA | 19684585 | NA | NA | NA | NA |
| MOBOCERTINIB | CONJUNCTIVITIS | Mobocertinib / EXKIVITY | 600 | 19405 | 2 | 19684585 | 3.381605078 | 598 | 19403 | 19664582 |
| MOBOCERTINIB | CORNEAL EPITHELIAL DEFECT | Mobocertinib / EXKIVITY | 600 | 99 | NA | 19684585 | NA | NA | NA | NA |
| MOBOCERTINIB | CORNEAL EROSION | Mobocertinib / EXKIVITY | 600 | 496 | NA | 19684585 | NA | NA | NA | NA |
| MOBOCERTINIB | CORNEAL MELT | Mobocertinib / EXKIVITY | 600 | -Inf | NA | 19684585 | NA | NA | NA | NA |
| MOBOCERTINIB | CORNEAL PERFORATION | Mobocertinib / EXKIVITY | 600 | 576 | NA | 19684585 | NA | NA | NA | NA |
| MOBOCERTINIB | CORNEAL ULCER | Mobocertinib / EXKIVITY | 600 | 101 | NA | 19684585 | NA | NA | NA | NA |
| MOBOCERTINIB | CORNEAL ULCERATION | Mobocertinib / EXKIVITY | 600 | -Inf | NA | 19684585 | NA | NA | NA | NA |
| MOBOCERTINIB | CORNEAL VERTICILLATA | Mobocertinib / EXKIVITY | 600 | -Inf | NA | 19684585 | NA | NA | NA | NA |
| MOBOCERTINIB | DRY EYE | Mobocertinib / EXKIVITY | 600 | 43203 | 6 | 19684585 | 4.556794453 | 594 | 43197 | 19640788 |

| **Table S2.** Complete Signal Data | | | | | | | | | | |
|---|---|---|---|---|---|---|---|---|---|---|
| Drug | Event | Label | total_drug_reports | total_event_reports | a | total_faers | prr | b | c | d |
| MOBOCERTINIB | EYE PAIN | Mobocertinib / EXKIVITY | 600 | 51035 | NA | 19684585 | NA | NA | NA | NA |
| MOBOCERTINIB | IRIDOCYCLITIS | Mobocertinib / EXKIVITY | 600 | 2852 | NA | 19684585 | NA | NA | NA | NA |
| MOBOCERTINIB | KERATITIS | Mobocertinib / EXKIVITY | 600 | 6613 | NA | 19684585 | NA | NA | NA | NA |
| MOBOCERTINIB | KERATOCONJUNCTIVITIS | Mobocertinib / EXKIVITY | 600 | 459 | NA | 19684585 | NA | NA | NA | NA |
| MOBOCERTINIB | LID ECTROPION | Mobocertinib / EXKIVITY | 600 | -Inf | NA | 19684585 | NA | NA | NA | NA |
| MOBOCERTINIB | MEIBOMIAN GLAND DYSFUNCTION | Mobocertinib / EXKIVITY | 600 | 383 | NA | 19684585 | NA | NA | NA | NA |
| MOBOCERTINIB | OPTIC NEURITIS | Mobocertinib / EXKIVITY | 600 | 8787 | 1 | 19684585 | 3.733967866 | 599 | 8786 | 19675199 |
| MOBOCERTINIB | OPTIC NEUROPATHY | Mobocertinib / EXKIVITY | 600 | 2365 | NA | 19684585 | NA | NA | NA | NA |
| MOBOCERTINIB | PERSISTENT CORNEAL EPITHELIAL DEFECT | Mobocertinib / EXKIVITY | 600 | 99 | NA | 19684585 | NA | NA | NA | NA |
| MOBOCERTINIB | PHOTOPHOBIA | Mobocertinib / EXKIVITY | 600 | 17555 | 1 | 19684585 | 1.868898352 | 599 | 17554 | 19666431 |
| MOBOCERTINIB | RETINAL HEMORRHAGE | Mobocertinib / EXKIVITY | 600 | -Inf | NA | 19684585 | NA | NA | NA | NA |
| MOBOCERTINIB | SUBCONJUNCTIVAL HEMORRHAGE | Mobocertinib / EXKIVITY | 600 | -Inf | NA | 19684585 | NA | NA | NA | NA |
| MOBOCERTINIB | TRICHIASIS | Mobocertinib / EXKIVITY | 600 | 287 | NA | 19684585 | NA | NA | NA | NA |
| MOBOCERTINIB | TRICHOMEGALY | Mobocertinib / EXKIVITY | 600 | 89 | NA | 19684585 | NA | NA | NA | NA |
| MOBOCERTINIB | UVEITIS | Mobocertinib / EXKIVITY | 600 | 13175 | NA | 19684585 | NA | NA | NA | NA |
| MOBOCERTINIB | VISION LOSS | Mobocertinib / EXKIVITY | 600 | 193 | NA | 19684585 | NA | NA | NA | NA |
| MOBOCERTINIB | VISUAL ACUITY REDUCED | Mobocertinib / EXKIVITY | 600 | 32697 | NA | 19684585 | NA | NA | NA | NA |
| MOBOCERTINIB | VORTEX KERATOPATHY | Mobocertinib / EXKIVITY | 600 | -Inf | NA | 19684585 | NA | NA | NA | NA |
| CETUXIMAB | BLEPHARITIS | Cetuximab / Erbitux | 12520 | 3685 | 6 | 19684585 | 2.562518942 | 12514 | 3679 | 19668386 |
| CETUXIMAB | CONJUNCTIVAL HEMORRHAGE | Cetuximab / Erbitux | 12520 | -Inf | NA | 19684585 | NA | NA | NA | NA |
| CETUXIMAB | CONJUNCTIVITIS | Cetuximab / Erbitux | 12520 | 19405 | 49 | 19684585 | 3.977645624 | 12471 | 19356 | 19652709 |
| CETUXIMAB | CORNEAL EPITHELIAL DEFECT | Cetuximab / Erbitux | 12520 | 99 | NA | 19684585 | NA | NA | NA | NA |
| CETUXIMAB | CORNEAL EROSION | Cetuximab / Erbitux | 12520 | 496 | 1 | 19684585 | 3.174244845 | 12519 | 495 | 19671570 |
| CETUXIMAB | CORNEAL MELT | Cetuximab / Erbitux | 12520 | -Inf | NA | 19684585 | NA | NA | NA | NA |
| CETUXIMAB | CORNEAL PERFORATION | Cetuximab / Erbitux | 12520 | 576 | 1 | 19684585 | 2.732610779 | 12519 | 575 | 19671490 |
| CETUXIMAB | CORNEAL ULCER | Cetuximab / Erbitux | 12520 | 101 | NA | 19684585 | NA | NA | NA | NA |
| CETUXIMAB | CORNEAL ULCERATION | Cetuximab / Erbitux | 12520 | -Inf | NA | 19684585 | NA | NA | NA | NA |
| CETUXIMAB | CORNEAL VERTICILLATA | Cetuximab / Erbitux | 12520 | -Inf | NA | 19684585 | NA | NA | NA | NA |
| CETUXIMAB | DRY EYE | Cetuximab / Erbitux | 12520 | 43203 | 7 | 19684585 | 0.2546244649 | 12513 | 43196 | 19628869 |
| CETUXIMAB | EYE PAIN | Cetuximab / Erbitux | 12520 | 51035 | 1 | 19684585 | 0.03078832147 | 12519 | 51034 | 19621031 |
| CETUXIMAB | IRIDOCYCLITIS | Cetuximab / Erbitux | 12520 | 2852 | 1 | 19684585 | 0.5511228334 | 12519 | 2851 | 19669214 |
| CETUXIMAB | KERATITIS | Cetuximab / Erbitux | 12520 | 6613 | 18 | 19684585 | 4.288479388 | 12502 | 6595 | 19665470 |
| CETUXIMAB | KERATOCONJUNCTIVITIS | Cetuximab / Erbitux | 12520 | 459 | 3 | 19684585 | 10.33717893 | 12517 | 456 | 19671609 |
| CETUXIMAB | LID ECTROPION | Cetuximab / Erbitux | 12520 | -Inf | NA | 19684585 | NA | NA | NA | NA |
| CETUXIMAB | MEIBOMIAN GLAND DYSFUNCTION | Cetuximab / Erbitux | 12520 | 383 | NA | 19684585 | NA | NA | NA | NA |
| CETUXIMAB | OPTIC NEURITIS | Cetuximab / Erbitux | 12520 | 8787 | 5 | 19684585 | 0.894586198 | 12515 | 8782 | 19663283 |
| CETUXIMAB | OPTIC NEUROPATHY | Cetuximab / Erbitux | 12520 | 2365 | NA | 19684585 | NA | NA | NA | NA |
| CETUXIMAB | PERSISTENT CORNEAL EPITHELIAL DEFECT | Cetuximab / Erbitux | 12520 | 99 | NA | 19684585 | NA | NA | NA | NA |
| CETUXIMAB | PHOTOPHOBIA | Cetuximab / Erbitux | 12520 | 17555 | 1 | 19684585 | 0.08950958175 | 12519 | 17554 | 19654511 |
| CETUXIMAB | RETINAL HEMORRHAGE | Cetuximab / Erbitux | 12520 | -Inf | NA | 19684585 | NA | NA | NA | NA |
| CETUXIMAB | SUBCONJUNCTIVAL HEMORRHAGE | Cetuximab / Erbitux | 12520 | -Inf | NA | 19684585 | NA | NA | NA | NA |
| CETUXIMAB | TRICHIASIS | Cetuximab / Erbitux | 12520 | 287 | NA | 19684585 | NA | NA | NA | NA |
| CETUXIMAB | TRICHOMEGALY | Cetuximab / Erbitux | 12520 | 89 | 8 | 19684585 | 155.1853035 | 12512 | 81 | 19671984 |
| CETUXIMAB | UVEITIS | Cetuximab / Erbitux | 12520 | 13175 | 5 | 19684585 | 0.5965266508 | 12515 | 13170 | 19658895 |
| CETUXIMAB | VISION LOSS | Cetuximab / Erbitux | 12520 | 193 | 1 | 19684585 | 8.18359999 | 12519 | 192 | 19671873 |
| CETUXIMAB | VISUAL ACUITY REDUCED | Cetuximab / Erbitux | 12520 | 32697 | 11 | 19684585 | 0.5287818387 | 12509 | 32686 | 19639379 |
| CETUXIMAB | VORTEX KERATOPATHY | Cetuximab / Erbitux | 12520 | -Inf | NA | 19684585 | NA | NA | NA | NA |
| PANITUMUMAB | BLEPHARITIS | Panitumumab / Vectibix | 5964 | 3685 | 9 | 19684585 | 8.078375449 | 5955 | 3676 | 19674945 |
| PANITUMUMAB | CONJUNCTIVAL HEMORRHAGE | Panitumumab / Vectibix | 5964 | -Inf | NA | 19684585 | NA | NA | NA | NA |
| PANITUMUMAB | CONJUNCTIVITIS | Panitumumab / Vectibix | 5964 | 19405 | 54 | 19684585 | 9.207619704 | 5910 | 19351 | 19659270 |
| PANITUMUMAB | CORNEAL EPITHELIAL DEFECT | Panitumumab / Vectibix | 5964 | 99 | NA | 19684585 | NA | NA | NA | NA |
| PANITUMUMAB | CORNEAL EROSION | Panitumumab / Vectibix | 5964 | 496 | NA | 19684585 | NA | NA | NA | NA |

| **Table S2.** Complete Signal Data | | | | | | | | | | |
|---|---|---|---|---|---|---|---|---|---|---|
| Drug | Event | Label | total_drug_reports | total_event_reports | a | total_faers | prr | b | c | d |
| PANITUMUMAB | CORNEAL MELT | Panitumumab / Vectibix | 5964 | -Inf | NA | 19684585 | NA | NA | NA | NA |
| PANITUMUMAB | CORNEAL PERFORATION | Panitumumab / Vectibix | 5964 | 576 | NA | 19684585 | NA | NA | NA | NA |
| PANITUMUMAB | CORNEAL ULCER | Panitumumab / Vectibix | 5964 | 101 | NA | 19684585 | NA | NA | NA | NA |
| PANITUMUMAB | CORNEAL ULCERATION | Panitumumab / Vectibix | 5964 | -Inf | NA | 19684585 | NA | NA | NA | NA |
| PANITUMUMAB | CORNEAL VERTICILLATA | Panitumumab / Vectibix | 5964 | -Inf | NA | 19684585 | NA | NA | NA | NA |
| PANITUMUMAB | DRY EYE | Panitumumab / Vectibix | 5964 | 43203 | 13 | 19684585 | 0.9931553239 | 5951 | 43190 | 19635431 |
| PANITUMUMAB | EYE PAIN | Panitumumab / Vectibix | 5964 | 51035 | 1 | 19684585 | 0.06465430051 | 5963 | 51034 | 19627587 |
| PANITUMUMAB | IRIDOCYCLITIS | Panitumumab / Vectibix | 5964 | 2852 | 3 | 19684585 | 3.474448128 | 5961 | 2849 | 19675772 |
| PANITUMUMAB | KERATITIS | Panitumumab / Vectibix | 5964 | 6613 | 9 | 19684585 | 4.496685062 | 5955 | 6604 | 19672017 |
| PANITUMUMAB | KERATOCONJUNCTIVITIS | Panitumumab / Vectibix | 5964 | 459 | NA | 19684585 | NA | NA | NA | NA |
| PANITUMUMAB | LID ECTROPION | Panitumumab / Vectibix | 5964 | -Inf | NA | 19684585 | NA | NA | NA | NA |
| PANITUMUMAB | MEIBOMIAN GLAND DYSFUNCTION | Panitumumab / Vectibix | 5964 | 383 | NA | 19684585 | NA | NA | NA | NA |
| PANITUMUMAB | OPTIC NEURITIS | Panitumumab / Vectibix | 5964 | 8787 | NA | 19684585 | NA | NA | NA | NA |
| PANITUMUMAB | OPTIC NEUROPATHY | Panitumumab / Vectibix | 5964 | 2365 | 1 | 19684585 | 1.395756164 | 5963 | 2364 | 19676257 |
| PANITUMUMAB | PERSISTENT CORNEAL EPITHELIAL DEFECT | Panitumumab / Vectibix | 5964 | 99 | NA | 19684585 | NA | NA | NA | NA |
| PANITUMUMAB | PHOTOPHOBIA | Panitumumab / Vectibix | 5964 | 17555 | 10 | 19684585 | 1.880631275 | 5954 | 17545 | 19661076 |
| PANITUMUMAB | RETINAL HEMORRHAGE | Panitumumab / Vectibix | 5964 | -Inf | NA | 19684585 | NA | NA | NA | NA |
| PANITUMUMAB | SUBCONJUNCTIVAL HEMORRHAGE | Panitumumab / Vectibix | 5964 | -Inf | NA | 19684585 | NA | NA | NA | NA |
| PANITUMUMAB | TRICHIASIS | Panitumumab / Vectibix | 5964 | 287 | 1 | 19684585 | 11.53694955 | 5963 | 286 | 19678335 |
| PANITUMUMAB | TRICHOMEGALY | Panitumumab / Vectibix | 5964 | 89 | 11 | 19684585 | 465.323632 | 5953 | 78 | 19678543 |
| PANITUMUMAB | UVEITIS | Panitumumab / Vectibix | 5964 | 13175 | 5 | 19684585 | 1.252683209 | 5959 | 13170 | 19665451 |
| PANITUMUMAB | VISION LOSS | Panitumumab / Vectibix | 5964 | 193 | NA | 19684585 | NA | NA | NA | NA |
| PANITUMUMAB | VISUAL ACUITY REDUCED | Panitumumab / Vectibix | 5964 | 32697 | 5 | 19684585 | 0.5046444959 | 5959 | 32692 | 19645929 |
| PANITUMUMAB | VORTEX KERATOPATHY | Panitumumab / Vectibix | 5964 | -Inf | NA | 19684585 | NA | NA | NA | NA |
| NECITUMUMAB | BLEPHARITIS | Necitumumab / Portrazza | 475 | 3685 | NA | 19684585 | NA | NA | NA | NA |
| NECITUMUMAB | CONJUNCTIVAL HEMORRHAGE | Necitumumab / Portrazza | 475 | -Inf | NA | 19684585 | NA | NA | NA | NA |
| NECITUMUMAB | CONJUNCTIVITIS | Necitumumab / Portrazza | 475 | 19405 | 1 | 19684585 | 2.13565407 | 474 | 19404 | 19664706 |
| NECITUMUMAB | CORNEAL EPITHELIAL DEFECT | Necitumumab / Portrazza | 475 | 99 | NA | 19684585 | NA | NA | NA | NA |
| NECITUMUMAB | CORNEAL EROSION | Necitumumab / Portrazza | 475 | 496 | NA | 19684585 | NA | NA | NA | NA |
| NECITUMUMAB | CORNEAL MELT | Necitumumab / Portrazza | 475 | -Inf | NA | 19684585 | NA | NA | NA | NA |
| NECITUMUMAB | CORNEAL PERFORATION | Necitumumab / Portrazza | 475 | 576 | NA | 19684585 | NA | NA | NA | NA |
| NECITUMUMAB | CORNEAL ULCER | Necitumumab / Portrazza | 475 | 101 | NA | 19684585 | NA | NA | NA | NA |
| NECITUMUMAB | CORNEAL ULCERATION | Necitumumab / Portrazza | 475 | -Inf | NA | 19684585 | NA | NA | NA | NA |
| NECITUMUMAB | CORNEAL VERTICILLATA | Necitumumab / Portrazza | 475 | -Inf | NA | 19684585 | NA | NA | NA | NA |
| NECITUMUMAB | DRY EYE | Necitumumab / Portrazza | 475 | 43203 | NA | 19684585 | NA | NA | NA | NA |
| NECITUMUMAB | EYE PAIN | Necitumumab / Portrazza | 475 | 51035 | NA | 19684585 | NA | NA | NA | NA |
| NECITUMUMAB | IRIDOCYCLITIS | Necitumumab / Portrazza | 475 | 2852 | NA | 19684585 | NA | NA | NA | NA |
| NECITUMUMAB | KERATITIS | Necitumumab / Portrazza | 475 | 6613 | NA | 19684585 | NA | NA | NA | NA |
| NECITUMUMAB | KERATOCONJUNCTIVITIS | Necitumumab / Portrazza | 475 | 459 | NA | 19684585 | NA | NA | NA | NA |
| NECITUMUMAB | LID ECTROPION | Necitumumab / Portrazza | 475 | -Inf | NA | 19684585 | NA | NA | NA | NA |
| NECITUMUMAB | MEIBOMIAN GLAND DYSFUNCTION | Necitumumab / Portrazza | 475 | 383 | NA | 19684585 | NA | NA | NA | NA |
| NECITUMUMAB | OPTIC NEURITIS | Necitumumab / Portrazza | 475 | 8787 | NA | 19684585 | NA | NA | NA | NA |
| NECITUMUMAB | OPTIC NEUROPATHY | Necitumumab / Portrazza | 475 | 2365 | NA | 19684585 | NA | NA | NA | NA |
| NECITUMUMAB | PERSISTENT CORNEAL EPITHELIAL DEFECT | Necitumumab / Portrazza | 475 | 99 | NA | 19684585 | NA | NA | NA | NA |
| NECITUMUMAB | PHOTOPHOBIA | Necitumumab / Portrazza | 475 | 17555 | NA | 19684585 | NA | NA | NA | NA |
| NECITUMUMAB | RETINAL HEMORRHAGE | Necitumumab / Portrazza | 475 | -Inf | NA | 19684585 | NA | NA | NA | NA |
| NECITUMUMAB | SUBCONJUNCTIVAL HEMORRHAGE | Necitumumab / Portrazza | 475 | -Inf | NA | 19684585 | NA | NA | NA | NA |
| NECITUMUMAB | TRICHIASIS | Necitumumab / Portrazza | 475 | 287 | NA | 19684585 | NA | NA | NA | NA |
| NECITUMUMAB | TRICHOMEGALY | Necitumumab / Portrazza | 475 | 89 | NA | 19684585 | NA | NA | NA | NA |
| NECITUMUMAB | UVEITIS | Necitumumab / Portrazza | 475 | 13175 | NA | 19684585 | NA | NA | NA | NA |
| NECITUMUMAB | VISION LOSS | Necitumumab / Portrazza | 475 | 193 | NA | 19684585 | NA | NA | NA | NA |
| NECITUMUMAB | VISUAL ACUITY REDUCED | Necitumumab / Portrazza | 475 | 32697 | NA | 19684585 | NA | NA | NA | NA |

| **Table S2.** Complete Signal Data | | | | | | | | | | |
|---|---|---|---|---|---|---|---|---|---|---|
| Drug | Event | Label | total_drug_reports | total_event_reports | a | total_faers | prr | b | c | d |
| NECITUMUMAB | VORTEX KERATOPATHY | Necitumumab / Portrazza | 475 | -Inf | NA | 19684585 | NA | NA | NA | NA |
| VANDETANIB | BLEPHARITIS | Vandetanib | 729 | 3685 | NA | 19684585 | NA | NA | NA | NA |
| VANDETANIB | CONJUNCTIVAL HEMORRHAGE | Vandetanib | 729 | -Inf | NA | 19684585 | NA | NA | NA | NA |
| VANDETANIB | CONJUNCTIVITIS | Vandetanib | 729 | 19405 | NA | 19684585 | NA | NA | NA | NA |
| VANDETANIB | CORNEAL EPITHELIAL DEFECT | Vandetanib | 729 | 99 | NA | 19684585 | NA | NA | NA | NA |
| VANDETANIB | CORNEAL EROSION | Vandetanib | 729 | 496 | NA | 19684585 | NA | NA | NA | NA |
| VANDETANIB | CORNEAL MELT | Vandetanib | 729 | -Inf | NA | 19684585 | NA | NA | NA | NA |
| VANDETANIB | CORNEAL PERFORATION | Vandetanib | 729 | 576 | NA | 19684585 | NA | NA | NA | NA |
| VANDETANIB | CORNEAL ULCER | Vandetanib | 729 | 101 | NA | 19684585 | NA | NA | NA | NA |
| VANDETANIB | CORNEAL ULCERATION | Vandetanib | 729 | -Inf | NA | 19684585 | NA | NA | NA | NA |
| VANDETANIB | CORNEAL VERTICILLATA | Vandetanib | 729 | -Inf | NA | 19684585 | NA | NA | NA | NA |
| VANDETANIB | DRY EYE | Vandetanib | 729 | 43203 | NA | 19684585 | NA | NA | NA | NA |
| VANDETANIB | EYE PAIN | Vandetanib | 729 | 51035 | NA | 19684585 | NA | NA | NA | NA |
| VANDETANIB | IRIDOCYCLITIS | Vandetanib | 729 | 2852 | NA | 19684585 | NA | NA | NA | NA |
| VANDETANIB | KERATITIS | Vandetanib | 729 | 6613 | NA | 19684585 | NA | NA | NA | NA |
| VANDETANIB | KERATOCONJUNCTIVITIS | Vandetanib | 729 | 459 | NA | 19684585 | NA | NA | NA | NA |
| VANDETANIB | LID ECTROPION | Vandetanib | 729 | -Inf | NA | 19684585 | NA | NA | NA | NA |
| VANDETANIB | MEIBOMIAN GLAND DYSFUNCTION | Vandetanib | 729 | 383 | 1 | 19684585 | 70.68370212 | 728 | 382 | 19683474 |
| VANDETANIB | OPTIC NEURITIS | Vandetanib | 729 | 8787 | NA | 19684585 | NA | NA | NA | NA |
| VANDETANIB | OPTIC NEUROPATHY | Vandetanib | 729 | 2365 | NA | 19684585 | NA | NA | NA | NA |
| VANDETANIB | PERSISTENT CORNEAL EPITHELIAL DEFECT | Vandetanib | 729 | 99 | NA | 19684585 | NA | NA | NA | NA |
| VANDETANIB | PHOTOPHOBIA | Vandetanib | 729 | 17555 | 1 | 19684585 | 1.538177863 | 728 | 17554 | 19666302 |
| VANDETANIB | RETINAL HEMORRHAGE | Vandetanib | 729 | -Inf | NA | 19684585 | NA | NA | NA | NA |
| VANDETANIB | SUBCONJUNCTIVAL HEMORRHAGE | Vandetanib | 729 | -Inf | NA | 19684585 | NA | NA | NA | NA |
| VANDETANIB | TRICHIASIS | Vandetanib | 729 | 287 | NA | 19684585 | NA | NA | NA | NA |
| VANDETANIB | TRICHOMEGALY | Vandetanib | 729 | 89 | NA | 19684585 | NA | NA | NA | NA |
| VANDETANIB | UVEITIS | Vandetanib | 729 | 13175 | NA | 19684585 | NA | NA | NA | NA |
| VANDETANIB | VISION LOSS | Vandetanib | 729 | 193 | NA | 19684585 | NA | NA | NA | NA |
| VANDETANIB | VISUAL ACUITY REDUCED | Vandetanib | 729 | 32697 | 2 | 19684585 | 1.651700518 | 727 | 32695 | 19651161 |
| VANDETANIB | VORTEX KERATOPATHY | Vandetanib | 729 | -Inf | NA | 19684585 | NA | NA | NA | NA |
| AMIVANTAMAB | BLEPHARITIS | Amivantamab / Rybrevant | 783 | 3685 | NA | 19684585 | NA | NA | NA | NA |
| AMIVANTAMAB | CONJUNCTIVAL HEMORRHAGE | Amivantamab / Rybrevant | 783 | -Inf | NA | 19684585 | NA | NA | NA | NA |
| AMIVANTAMAB | CONJUNCTIVITIS | Amivantamab / Rybrevant | 783 | 19405 | 3 | 19684585 | 3.887066586 | 780 | 19402 | 19664400 |
| AMIVANTAMAB | CORNEAL EPITHELIAL DEFECT | Amivantamab / Rybrevant | 783 | 99 | NA | 19684585 | NA | NA | NA | NA |
| AMIVANTAMAB | CORNEAL EROSION | Amivantamab / Rybrevant | 783 | 496 | 1 | 19684585 | 50.78576828 | 782 | 495 | 19683307 |
| AMIVANTAMAB | CORNEAL MELT | Amivantamab / Rybrevant | 783 | -Inf | NA | 19684585 | NA | NA | NA | NA |
| AMIVANTAMAB | CORNEAL PERFORATION | Amivantamab / Rybrevant | 783 | 576 | NA | 19684585 | NA | NA | NA | NA |
| AMIVANTAMAB | CORNEAL ULCER | Amivantamab / Rybrevant | 783 | 101 | NA | 19684585 | NA | NA | NA | NA |
| AMIVANTAMAB | CORNEAL ULCERATION | Amivantamab / Rybrevant | 783 | -Inf | NA | 19684585 | NA | NA | NA | NA |
| AMIVANTAMAB | CORNEAL VERTICILLATA | Amivantamab / Rybrevant | 783 | -Inf | NA | 19684585 | NA | NA | NA | NA |
| AMIVANTAMAB | DRY EYE | Amivantamab / Rybrevant | 783 | 43203 | NA | 19684585 | NA | NA | NA | NA |
| AMIVANTAMAB | EYE PAIN | Amivantamab / Rybrevant | 783 | 51035 | NA | 19684585 | NA | NA | NA | NA |
| AMIVANTAMAB | IRIDOCYCLITIS | Amivantamab / Rybrevant | 783 | 2852 | NA | 19684585 | NA | NA | NA | NA |
| AMIVANTAMAB | KERATITIS | Amivantamab / Rybrevant | 783 | 6613 | 4 | 19684585 | 15.21498278 | 779 | 6609 | 19677193 |
| AMIVANTAMAB | KERATOCONJUNCTIVITIS | Amivantamab / Rybrevant | 783 | 459 | NA | 19684585 | NA | NA | NA | NA |
| AMIVANTAMAB | LID ECTROPION | Amivantamab / Rybrevant | 783 | -Inf | NA | 19684585 | NA | NA | NA | NA |
| AMIVANTAMAB | MEIBOMIAN GLAND DYSFUNCTION | Amivantamab / Rybrevant | 783 | 383 | NA | 19684585 | NA | NA | NA | NA |
| AMIVANTAMAB | OPTIC NEURITIS | Amivantamab / Rybrevant | 783 | 8787 | NA | 19684585 | NA | NA | NA | NA |
| AMIVANTAMAB | OPTIC NEUROPATHY | Amivantamab / Rybrevant | 783 | 2365 | NA | 19684585 | NA | NA | NA | NA |
| AMIVANTAMAB | PERSISTENT CORNEAL EPITHELIAL DEFECT | Amivantamab / Rybrevant | 783 | 99 | NA | 19684585 | NA | NA | NA | NA |
| AMIVANTAMAB | PHOTOPHOBIA | Amivantamab / Rybrevant | 783 | 17555 | NA | 19684585 | NA | NA | NA | NA |
| AMIVANTAMAB | RETINAL HEMORRHAGE | Amivantamab / Rybrevant | 783 | -Inf | NA | 19684585 | NA | NA | NA | NA |

| **Table S2.** Complete Signal Data | | | | | | | | | | |
|---|---|---|---|---|---|---|---|---|---|---|
| Drug | Event | Label | total_drug_reports | total_event_reports | a | total_faers | prr | b | c | d |
| AMIVANTAMAB | SUBCONJUNCTIVAL HEMORRHAGE | Amivantamab / Rybrevant | 783 | -Inf | NA | 19684585 | NA | NA | NA | NA |
| AMIVANTAMAB | TRICHIASIS | Amivantamab / Rybrevant | 783 | 287 | NA | 19684585 | NA | NA | NA | NA |
| AMIVANTAMAB | TRICHOMEGALY | Amivantamab / Rybrevant | 783 | 89 | 1 | 19684585 | 285.6699466 | 782 | 88 | 19683714 |
| AMIVANTAMAB | UVEITIS | Amivantamab / Rybrevant | 783 | 13175 | NA | 19684585 | NA | NA | NA | NA |
| AMIVANTAMAB | VISION LOSS | Amivantamab / Rybrevant | 783 | 193 | NA | 19684585 | NA | NA | NA | NA |
| AMIVANTAMAB | VISUAL ACUITY REDUCED | Amivantamab / Rybrevant | 783 | 32697 | NA | 19684585 | NA | NA | NA | NA |
| AMIVANTAMAB | VORTEX KERATOPATHY | Amivantamab / Rybrevant | 783 | -Inf | NA | 19684585 | NA | NA | NA | NA |
| DEPATUXIZUMAB MAFODOTIN | BLEPHARITIS | Depatuxizumab mafodotin / ABT-414 | 13 | 3685 | NA | 19684585 | NA | NA | NA | NA |
| DEPATUXIZUMAB MAFODOTIN | CONJUNCTIVAL HEMORRHAGE | Depatuxizumab mafodotin / ABT-414 | 13 | -Inf | NA | 19684585 | NA | NA | NA | NA |
| DEPATUXIZUMAB MAFODOTIN | CONJUNCTIVITIS | Depatuxizumab mafodotin / ABT-414 | 13 | 19405 | NA | 19684585 | NA | NA | NA | NA |
| DEPATUXIZUMAB MAFODOTIN | CORNEAL EPITHELIAL DEFECT | Depatuxizumab mafodotin / ABT-414 | 13 | 99 | NA | 19684585 | NA | NA | NA | NA |
| DEPATUXIZUMAB MAFODOTIN | CORNEAL EROSION | Depatuxizumab mafodotin / ABT-414 | 13 | 496 | NA | 19684585 | NA | NA | NA | NA |
| DEPATUXIZUMAB MAFODOTIN | CORNEAL MELT | Depatuxizumab mafodotin / ABT-414 | 13 | -Inf | NA | 19684585 | NA | NA | NA | NA |
| DEPATUXIZUMAB MAFODOTIN | CORNEAL PERFORATION | Depatuxizumab mafodotin / ABT-414 | 13 | 576 | NA | 19684585 | NA | NA | NA | NA |
| DEPATUXIZUMAB MAFODOTIN | CORNEAL ULCER | Depatuxizumab mafodotin / ABT-414 | 13 | 101 | NA | 19684585 | NA | NA | NA | NA |
| DEPATUXIZUMAB MAFODOTIN | CORNEAL ULCERATION | Depatuxizumab mafodotin / ABT-414 | 13 | -Inf | NA | 19684585 | NA | NA | NA | NA |
| DEPATUXIZUMAB MAFODOTIN | CORNEAL VERTICILLATA | Depatuxizumab mafodotin / ABT-414 | 13 | -Inf | NA | 19684585 | NA | NA | NA | NA |
| DEPATUXIZUMAB MAFODOTIN | DRY EYE | Depatuxizumab mafodotin / ABT-414 | 13 | 43203 | NA | 19684585 | NA | NA | NA | NA |
| DEPATUXIZUMAB MAFODOTIN | EYE PAIN | Depatuxizumab mafodotin / ABT-414 | 13 | 51035 | NA | 19684585 | NA | NA | NA | NA |
| DEPATUXIZUMAB MAFODOTIN | IRIDOCYCLITIS | Depatuxizumab mafodotin / ABT-414 | 13 | 2852 | NA | 19684585 | NA | NA | NA | NA |
| DEPATUXIZUMAB MAFODOTIN | KERATITIS | Depatuxizumab mafodotin / ABT-414 | 13 | 6613 | NA | 19684585 | NA | NA | NA | NA |
| DEPATUXIZUMAB MAFODOTIN | KERATOCONJUNCTIVITIS | Depatuxizumab mafodotin / ABT-414 | 13 | 459 | NA | 19684585 | NA | NA | NA | NA |
| DEPATUXIZUMAB MAFODOTIN | LID ECTROPION | Depatuxizumab mafodotin / ABT-414 | 13 | -Inf | NA | 19684585 | NA | NA | NA | NA |
| DEPATUXIZUMAB MAFODOTIN | MEIBOMIAN GLAND DYSFUNCTION | Depatuxizumab mafodotin / ABT-414 | 13 | 383 | NA | 19684585 | NA | NA | NA | NA |
| DEPATUXIZUMAB MAFODOTIN | OPTIC NEURITIS | Depatuxizumab mafodotin / ABT-414 | 13 | 8787 | NA | 19684585 | NA | NA | NA | NA |
| DEPATUXIZUMAB MAFODOTIN | OPTIC NEUROPATHY | Depatuxizumab mafodotin / ABT-414 | 13 | 2365 | NA | 19684585 | NA | NA | NA | NA |
| DEPATUXIZUMAB MAFODOTIN | PERSISTENT CORNEAL EPITHELIAL DEFECT | Depatuxizumab mafodotin / ABT-414 | 13 | 99 | NA | 19684585 | NA | NA | NA | NA |
| DEPATUXIZUMAB MAFODOTIN | PHOTOPHOBIA | Depatuxizumab mafodotin / ABT-414 | 13 | 17555 | NA | 19684585 | NA | NA | NA | NA |
| DEPATUXIZUMAB MAFODOTIN | RETINAL HEMORRHAGE | Depatuxizumab mafodotin / ABT-414 | 13 | -Inf | NA | 19684585 | NA | NA | NA | NA |
| DEPATUXIZUMAB MAFODOTIN | SUBCONJUNCTIVAL HEMORRHAGE | Depatuxizumab mafodotin / ABT-414 | 13 | -Inf | NA | 19684585 | NA | NA | NA | NA |
| DEPATUXIZUMAB MAFODOTIN | TRICHIASIS | Depatuxizumab mafodotin / ABT-414 | 13 | 287 | NA | 19684585 | NA | NA | NA | NA |
| DEPATUXIZUMAB MAFODOTIN | TRICHOMEGALY | Depatuxizumab mafodotin / ABT-414 | 13 | 89 | NA | 19684585 | NA | NA | NA | NA |
| DEPATUXIZUMAB MAFODOTIN | UVEITIS | Depatuxizumab mafodotin / ABT-414 | 13 | 13175 | NA | 19684585 | NA | NA | NA | NA |
| DEPATUXIZUMAB MAFODOTIN | VISION LOSS | Depatuxizumab mafodotin / ABT-414 | 13 | 193 | NA | 19684585 | NA | NA | NA | NA |
| DEPATUXIZUMAB MAFODOTIN | VISUAL ACUITY REDUCED | Depatuxizumab mafodotin / ABT-414 | 13 | 32697 | NA | 19684585 | NA | NA | NA | NA |
| DEPATUXIZUMAB MAFODOTIN | VORTEX KERATOPATHY | Depatuxizumab mafodotin / ABT-414 | 13 | -Inf | NA | 19684585 | NA | NA | NA | NA |
